# The Prevalence, Risk Factors, and Antimicrobial Resistance of *Campylobacter* in African Children: A Systematic Review and Meta-Analysis

**DOI:** 10.64898/2026.02.09.26345948

**Authors:** Jordana Burdon Bailey, Alfred Menyere, Hannah Dawood, Oliver Mapila, Shareef Ngunguni, Gina Pinchbeck, Nicola Williams, Nigel Cunliffe, Jen Cornick

## Abstract

**Background:** *Campylobacter* is a major cause of childhood diarrhoea across Africa and asymptomatic carriage is frequently reported, however risk factors for *Campylobacter* presence remain poorly defined. This meta-analysis aimed to calculate the pooled prevalence of *Campylobacter* in diarrhoeic and non-diarrhoeic stool, assess its association with diarrhoea, identify risk factors for *Campylobacter* presence and antimicrobial resistance (AMR) patterns.

**Method:** English language studies on *Campylobacter* in children (<18 years) in Africa were searched. Prevalence of *Campylobacter* and AMR, Odds Ratios (OR) for *Campylobacter* presence in diarrhoeic stool and risk factors for *Campylobacter* were estimated. Heterogeneity was assessed using I^2^ and bias assessed via funnel plots and Egger’s test.

**Results:** A total of 168 studies were included in the meta-analysis. The pooled prevalence of *Campylobacter* in diarrhoeic stool was 11.25% (9.41-13.23%), in non-diarrhoeic stool 12.56% (7.79-18.27%), and mixed stool types 33.47% (20.53-47.81%). The OR for *Campylobacter* presence in diarrhoeic stool versus non-diarrhoeic stool was 1.95 (95% CI: 1.62-2.33). Age affected the OR with children 0-6 months old having an OR 2.57 (1.74-3.81), 7-12 months old an OR 1.60 (1.07-2.40), 13-24 months old an OR 1.02 (0.68-1.52) and 25-60 months old an OR 1.76 (0.77-4.05). Risk factors for *Campylobacter* presence in stool were children living in rural areas (pooled Adjusted Odds Ratio (pAOR) = 2.59 95% CI 1.43-4.69) and having contact with animals (pAOR 4.28 95% CI: 2.40-7.61). AMR prevalence ranged from 54.85% for ampicillin to 9.85% for chloramphenicol. Heterogeneity was high across all analyses.

**Conclusion:** *Campylobacter* prevalence is high in symptomatic and asymptomatic children across Africa. Contact with animals and living in an urban environment are risk factors for *Campylobacter* presence. Risk factor identification in the African context would be strengthened with standardized risk factors. Further research is needed to clarify the public health significance of asymptomatic carriage.

**What is already known on this topic –** *Campylobacter* is a significant cause of diarrhoea in children and asymptomatic carriage is common. However, the burden of asymptomatic carriage and risk factors are not well understood in Africa.

**What this study adds –** This meta-analysis highlights the high burden of asymptomatic *Campylobacter* carriage, its relation to age, and identified risk factors for *Campylobacter* in children in Africa.

**How this study might affect research, practice or policy –** Standardising risk factor assessments can guide future control strategies. Further research into the impact of asymptomatic carriage is warranted.

## INTRODUCTION

Gastrointestinal infections remain a major cause of child mortality (1) and morbidity, negatively impacting physical and intellectual development (2, 3). In particular, diarrhoea is a significant contributor to mortality in children under 5 years old (4). *Campylobacter* is a leading cause of gastroenteritis globally (5), and a significant contributor to childhood diarrhoea in Low– and Middle-Income Countries (LMICs) (6).

Until recently, research on *Campylobacter* has focused on High Income Countries (HICs) (7). In HICs, *Campylobacter* infection is predominantly foodborne and is strongly associated with the consumption of, or handling of chicken meat (8, 9). In contrast, studies from Africa suggest breastfeeding is associated with increased odds of *Campylobacter* colonization (10). These differences underscore the need for context-specific research to better define the epidemiology of *Campylobacter* in LMICs.

Campylobacteriosis is believed to be endemic in Africa, with repeated exposure leading to partially protective immunity (11). Asymptomatic carriage is common (12, 13) and has been linked to reduced height gain in Tanzanian children (14). This suggests asymptomatic *Campylobacter* carriage may impact child development.

Although campylobacteriosis is typically a self-limiting disease, antibiotic treatment is indicated in severe infections or immunocompromised individuals (15). Antimicrobial resistance (AMR), especially against first line treatments such as macrolides and fluoroquinolones, has been observed in *Campylobacter* (16). This is of particular concern in low resource settings where alternative antimicrobials may not be reliably available.

This review aimed to determine the prevalence, risk factors and AMR of *Campylobacter* in children in Africa. It investigated *Campylobacter* prevalence in children with diarrhoea and those without diarrhoea, to emphasize the extent of asymptomatic carriage. The findings of this study highlight risk factors relevant to the African context, providing a foundation for future research.

## METHODS

### Search Strategy

A systematic review and meta-analysis was conducted in accordance with the Preferred Reporting Items for Systematic Reviews and Meta-Analyses (PRISMA) guidelines (17). Medline, Web of Science and Scopus were searched for studies on 20^th^ August 2024 and updated on 5^th^ November 2025. Search terms and their related mesh terms included ‘*Campylobacter’,* ‘child’ and all African countries. The complete search strategy can be found in S1. To ensure inclusivity, country names included current and historical nomenclature. This review protocol was registered with Prospero (CRD42024582210).

### Eligibility criteria

Studies were included if they enrolled children (<18 years old). Any microbiological or molecular *Campylobacter* detection method and any country or countries within Africa were included. All observational studies and base-line data from intervention studies were eligible. Only literature published in English was included. There was no restriction on publication year.

Studies were excluded if they were systematic or literature reviews, letters, communications, book chapters, anecdotal evidence or from non-peer reviewed journals. Studies on oral *Campylobacter* and non-diarrhoeal sequelae of *Campylobacter* were excluded. Species historically linked to *Campylobacter*, such as *Helicobacter*, and studies on *Campylobacter* diarrhoea in people travelling to African countries were excluded.

### Study selection and data extraction

Search results were uploaded into Endnote and duplicates removed. Deduplicated literature was imported into Rayyan for screening (18). Screening of title and abstract, followed by full text screening, was done blind by two reviewers. Conflicts were discussed until a consensus was drawn, else a third reviewer was asked to adjudicate. Reasons for exclusion were recorded.

Data were extracted into Microsoft Excel by the primary reviewer and reviewed by a second reviewer. Disagreements were discussed and if necessary a third reviewer assessed the data. Authors were contacted where data were missing or for clarifications. If authors did not provide the requested information the data were excluded.

The following data were extracted: title, primary author, year of publication, journal, country/countries, setting (e.g. community or hospital), age group of participants, study type, sample type, method of confirming *Campylobacter*, number of samples tested, number of positives for *Campylobacter*, status of samples (diarrhoeic, non-diarrhoeic, or not differentiated referred to as ‘mixed’ in this review), species of *Campylobacter*, AMR, measurement e.g. odds ratios (OR).

Where several papers were published from the same study, each was assessed for eligibility. Where papers provided distinct datasets they were counted as separate papers, otherwise they were merged and a representative paper selected. Each paper was counted at abstract and full title screening, with ineligible papers discarded. Papers were grouped by study in the systematic review and meta-analysis.

### Risk of bias assessment

All eligible studies were assessed for bias and quality by two reviewers using the Joanna Briggs Institute (JBI) critical appraisal tool (19). Studies were assessed against their design type, however studies that calculated prevalence of *Campylobacter* only and therefore did not assess confounders were accounted for and included. Where responses to questions were “no” the reviewers discussed to determine if that affected the quality of the data extracted for the purpose of this review, and therefore whether it should be included or not. Where there were disagreements, these were discussed until resolved or a third reviewer consulted.

### Data analysis

All analyses were conducted in R using the meta and metafor packages, using a random effects model to account for between study heterogeneity. Publication bias was assessed via funnel plots and Egger’s test. Heterogeneity was assessed using Cochran’s Q statistic and Higgins’ I^2^ statistic. Where heterogeneity was high (I^2^ > 75% (20)), univariable meta-regression and subgroup analyses were conducted to investigate the source. The R^2^ statistic was used to assess moderator contribution towards heterogeneity in meta-regression.

Analyses were conducted at the genus level, with a separate pooled prevalence calculation for *C. jejuni* and *C. coli*. Meta-regression and subgroup analysis of age was done using the following categories: 0-6 months, 7-12 months, 13-24 months, 25-60 months and >60 months. Detection method was assessed using; a) the published method; b) methods categorised as culture, PCR, or other; and c) molecular or non-molecular. One method of detection was selected for studies utilising multiple methods, with culture preferentially selected. Study setting was assessed using a) published settings or b) grouped into community, clinic-based or not defined.

#### Prevalence

*Campylobacter* prevalence was extracted or calculated from cross-sectional studies, cohort studies and baseline data from randomized control studies. Pooled prevalence was calculated using random effects meta-analyses using inverse variance weighting and the Freeman-Tukey double arcsine transformation (21). The latter was used to stabilise outputs from studies with extreme prevalences.

Separate pooled prevalences were calculated for *Campylobacter spp., C. jejuni* and *C. coli* in normal, diarrhoeic and mixed stool types. Data for prevalence in mixed stool came from studies that did not contribute data towards prevalence analyses for diarrhoeic or non-diarrhoeic stool.

#### Odds Ratio for Campylobacter presence in diarrhoeic stool compared to non-diarrhoeic stool

Odds ratios (OR) were calculated from case-control, cross sectional and cohort studies that reported *Campylobacter* presence in diarrhoeic and non-diarrhoeic stools. To account for studies where no *Campylobacter* were detected in either group, a small continuity correction (+0.5) was added to all values. A random-effects meta-analysis was conducted on log transformed OR. Pooled effect estimates were back transformed for interpretation.

#### Odds Ratios for risk factors

Risk factors for *Campylobacter* presence in stool were investigated using OR. Crude and adjusted OR were analysed separately. Meta-analysis was conducted when at least 3 studies assessed the same risk factor. A random-effects meta-analysis was conducted on log transformed OR. Pooled effect estimates were back transformed for interpretation. Where meta-analysis identified influential studies, the analysis was repeated without those studies to determine their effect on the OR.

No studies assessed risk factors for *Campylobacter* presence in non-diarrhoeic stool. Due to the low number of studies investigating risk factors the meta-analysis was run on diarrhoeic and mixed stool types combined. Where 3 or more studies contributed data on *Campylobacter* presence in diarrhoeic stool, a separate meta-analysis was conducted.

#### Antimicrobial Resistance

Pooled AMR prevalence was calculated using random effects meta-analyses using inverse variance weighting and the Freeman-Tukey double arcsine transformation (21). Antibiotics assessed were those used traditionally as treatment for, or surveillance of AMR in *Campylobacter*. These included erythromycin (15μg), azithromycin (15μg), ciprofloxacin (5μg), doxycycline (30μg), tetracycline (30 μg), gentamicin (10μg), nalidixic acid (30μg), streptomycin (25μg) and chloramphenicol (30μg). The antibiotic concentrations noted above reflected the concentrations reported by Clinical and Laboratory Standards Institute (CLSI) as human clinical breakpoints for disk diffusion (22, 23). Studies that did not report MIC cut-offs against CLSI or European Committee on Antimicrobial Susceptibility Testing (EUCAST) were compared against values reported by CLSI (M45, 3^rd^ edition, 2016 for tetracycline, doxycycline, erythromycin and ciprofloxacin (23); M100, 30^th^ edition, 2020 for all others (22)). Intermediate resistance data were omitted from analysis, with total sample size adjusted for prevalence calculations. Meta-regression was conducted to assess sources of heterogeneity using decade, country and *Campylobacter* species as moderators.

### Patient and Public Involvement

As a systematic review and meta-analysis of previously published data patients and public were not involved in the design, conduct, reporting, or dissemination of this research.

## RESULTS

A total of 1,727 papers were identified with four additional papers added through citation searching. After removing duplicates (n=563) 1,168 papers were screened. At title/abstract screening 810 papers were removed, and 147 papers at full text assessment. Seven papers were removed after critical appraisal (S2), leaving 204 papers eligible for inclusion (10, 24–225) of which 168 contributed to the meta-analysis (See Fig 1).

**Fig 1.**
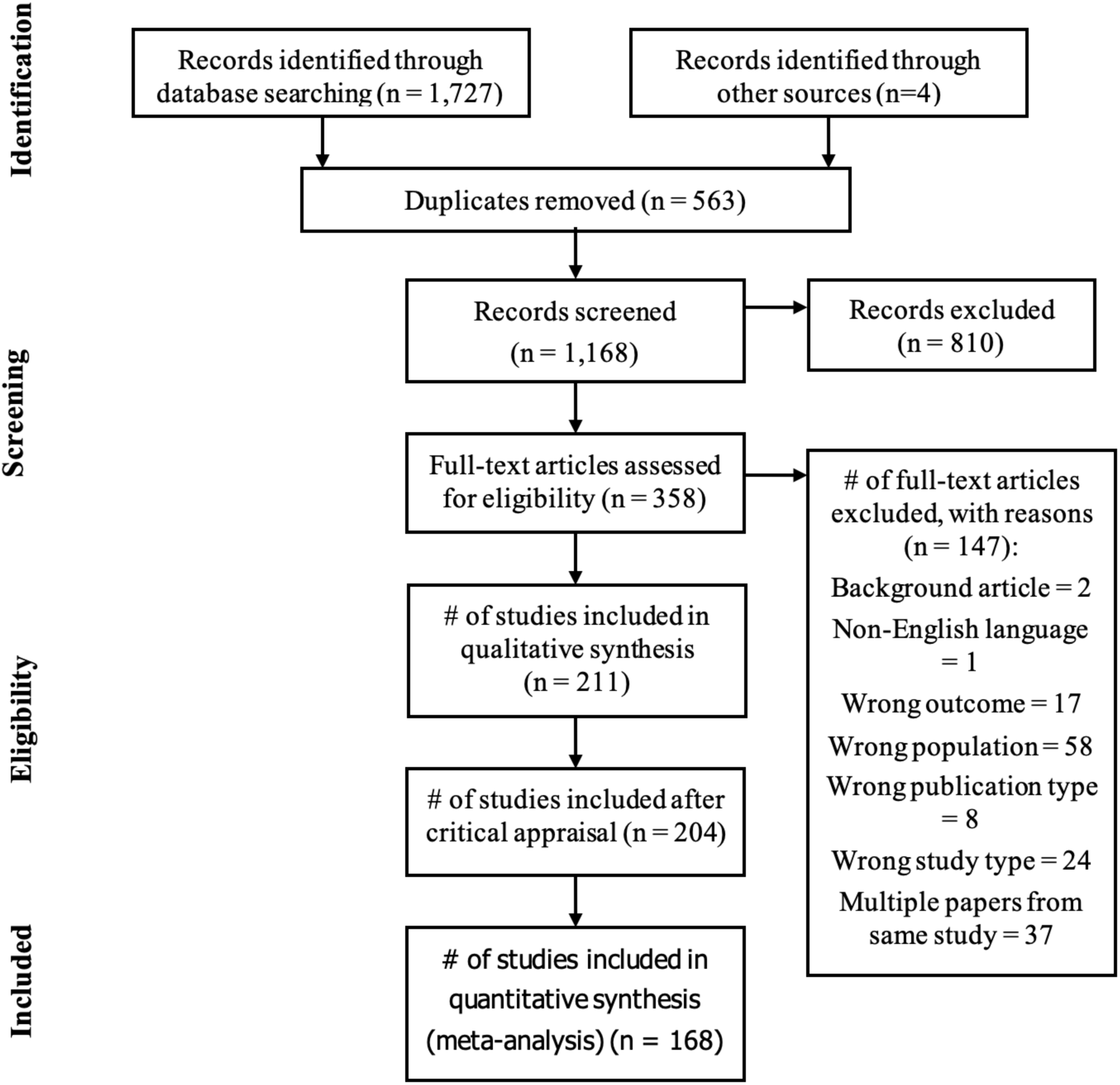
PRISMA flowchart for identification and inclusion of studies.

Papers included in the meta-analysis were published between 1979 and 2025 and included studies from 33 (61.11%) out of 54 African countries (S3). Egypt contributed the most papers (n = 23), followed by Ethiopia (n = 22), South Africa (n = 21), Kenya (n = 18), Tanzania (n = 14) and Nigeria (n = 15). All remaining countries contributed ≤five papers.

Most participants were recruited from a clinical setting (n = 125, 74.4%) (S3). Over two thirds of studies had participants whose maximum age was 60 months (n = 114, 67.86%) (S4). Studies identified *Campylobacter* mostly via culture (n = 130, 76.0%), followed by PCR (n = 34, 19.9%), with few studies using other methods, such as enzyme immunoassays measurements (S4). Over half of the papers were published since 2010 (n = 94, 55.95%) (S4), with a quarter of the studies published between 2020-2025 (S4). PCR contributed 26.42% (n = 14) and 45.45% (n = 20) of the total papers published in the decades 2010 and 2020 respectively. Culture was the main method of detection across all decades (S5).

### Pooled prevalence of *Campylobacter*

A summary of the pooled *Campylobacter* prevalences calculated using model-based calculations with weighted averages of back-transformed estimates is shown in Table 1. The highest *Campylobacter* prevalence was observed in mixed stool types (33.47%, 95% CI 20.53-47.81%). The prevalence in non-diarrhoeic stools (12.56%, 95% CI 7.79-18.27%) were slightly higher than in diarrhoeic stools (11.25%, 95% CI 9.41-13.23%). However, as the confidence intervals overlap and the interval for non-diarrheic stool is wider, the difference was not statistically significant.

**Table 1.** Pooled prevalence and meta-analysis for all *Campylobacter* spp., *C. jejuni and C. coli* in diarrhoeic stool, non-diarrhoeic stool and mixed (non-differentiated) stools.

|  | Diarrhoeic stools | Non-diarrhoeic stools | Mixed stools |
| --- | --- | --- | --- |
| *Campylobacter* spp |  |  |  |
| Number of studies | 108 | 32 | 15 |
| Model-based pooled prevalence and 95% CI | 11.25% (9.41-13.23) | 12.56% (7.79-18.27) | 33.47% (20.53-47.81%) |
| p-value for model | < .0001 | < .0001 | < .0001 |
| I<sup>2</sup> | 98.29% | 99.44% | 99.10% |
| H<sup>2</sup> | 58.36 | 177.76 | 110.5 |
| Q | (df = 107) = 4534 | (df = 31) = 4648 | (df = 14) = 1015 |
| Eggers test | z = 3.5761, p = 0.0003 | z = 0.0873, p = 0.9305 | z = 1.5450, p = 0.1223 |
| ***C. jejuni*** |  |  |  |
| Number of studies | 42 | 13 | 3 |
| Model-based pooled prevalence and 95% CI | 14.63% (9.68-20.39%) | 8.43% (1.83-19.05%) | 9.71% (3.37-18.76%) |
| p-value of model | < .0001 | < .0001 | < .0001 |
| I<sup>2</sup> | 98.81% | 99.17% | 88.27% |
| H<sup>2</sup> | 84.25 | 119.76 | 8.53 |
| Q | (df = 41) = 2372 | (df = 12) = 363 | (df = 2) = 21 |
| Eggers test | z = 1.3831, p = 0.1666 | z = 1.6724, p = 0.0944 | z = 4.6001, p < .0001 |
| ***C. coli*** |  |  |  |
| Number of studies | 23 | 6 | 3 |
| Model-based pooled prevalence and 95% CI | 5.40% (2.57-9.16%) | 2.82% (0.70-6.18%) | 2.55% (0.61-5.23%) |
| p-value for model | < .0001 | < .0001 | < .0001 |
| I<sup>2</sup> | 97.66% | 88.24% | 66.87% |
| H<sup>2</sup> | 42.81 | 8.5 | 3.02 |
| Q | (df = 22) = 972 | (df = 5) = 51 | (df = 2) = 6 |
| Eggers test | z = 1.7351, p = 0.0827 | z = 0.8586, p = 0.3906 | z = 1.1812, p = 0.2375 |

Heterogeneity was high across all pooled prevalence estimates (all I^2^ > 98%). Potential sources of heterogeneity were investigated through meta-regression. For *Campylobacter* prevalence in diarrhoeic stools, country contributed most towards heterogeneity (R^2^ = 22.30%). Decade, setting, study type, maximum age of participants, and comparing molecular to non-molecular methods did not explain any of the heterogeneity (R^2^ = 0.00% for all) (S6).

For non-diarrhoeic stools, one study was identified as a potential outlier. Removing this study and rerunning the metanalysis had little effect on the pooled prevalence, from 12.56% (95% CI: 7.79%-18.27%) to 11.20% (95% CI: 7.16-16.02%), though reduced it below the prevalence of diarrhoeic stool. Meta-regression was conducted on the full data set. Country explained the greatest amount of between-study heterogeneity (R^2^ = 62.70%), followed by decade (R^2^ = 59.53%) and detection method (R^2^ = 35.05). Setting and study type did not reduce heterogeneity (R^2^ = 0.0% for both). Heterogeneity remained high (I^2^ > 97%) for each moderator (S7).

For mixed stool type, moderators had minimal effect on reducing heterogeneity (I^2^ > 94%). Country accounted for most heterogeneity (R^2^ = 66.05%) followed by *Campylobacter* detection method type (R^2^ = 38.74%) and decade (R^2^ = 35.21%) (S8).

Publication bias was assessed using visual inspection of funnel plots and Egger’s test across all three stool categories. Egger’s tests did not provide significant evidence of bias or asymmetry for mixed stool (z = 1.5450, p = 0.1223) and non-diarrhoeic stool (z = 0.0873, p = 0.9305). There was a suggestion of publication bias or small study effects for diarrhoeic stools (z = 3.5761, p = 0.0003). Trim and fill analysis indicated that no studies were missing, indicating publication bias may be limited.

The pooled prevalence of *C. jejuni* and *C. coli,* the species mainly associated with diarrhoea, were calculated. Heterogeneity was high for all analyses (I^2^ > 88%) other than mixed stool type for *C. coli,* which was moderate (I^2^ = 66.87%). (Table 1). The pooled prevalence for *C. jejuni* was highest in diarrhoeic stool (14.63%, 95% CI: 9.68-20.39%) followed by mixed stool (9.71%, 95% CI: 3.37-18.76%) then non-diarrhoeic stool (8.43%, 95% CI:1.83-19.05%). The confidence intervals overlapped, suggesting these differences were not significant. The pooled prevalence of *C. coli* was less than *C. jejuni*, with diarrhoeic stool having the highest prevalence (5.40%: 95% CI: 2.57-9.16%) followed by non-diarrhoeic stool (2.82%, 95% CI: 0.70-6.18%) then mixed stool (2.55%, 95% CI: 0.61-5.23%). Confidence intervals overlapped suggesting the differences were not significant.

Pooled prevalences were calculated by stool type and method of detection (S9). The pooled prevalence in diarrhoeic stool varied from 10.89% (6.21-16.67%) for PCR to 21.61% (9.5-36.94%) for EIA. In non-diarrhoeic stool culture had the lowest prevalence (7.18%, 95%CI 4.04-11.11%) and PCR the highest (30.14%, 95% CI: 16.01-46.53). In mixed stool types PCR had the highest pooled prevalence (51.87%, 95% CI: 37.64-65.95%) with culture having a pooled prevalence of 20.41% (95% CI: 7.18-38.15%).

### Odds Ratio for *Campylobacter* presence in diarrhoeic versus non-diarrhoeic stool

Fifty-nine studies had data on *Campylobacter* presence in diarrhoeic and non-diarrhoeic stool, from which OR were calculated. Diarrhoeic stool had 1.95 times higher odds of having *Campylobacter* detected than non-diarrhoeic stool (OR = 1.95, 95% CI: 1.62-2.33, p < 0.0001). However, significant heterogeneity was observed across studies (I^2^ = 85.24%, p < 0.0001).

Visual inspection of the funnel plot and Egger’s regression suggested significant asymmetry (z = 3.6801, p = 0.0002). A trim and fill analysis indicated that 18 studies were potentially missing, suggesting publication bias. After adjustment, the OR for *Campylobacter* presence in diarrhoeic stool was 1.44 (95% CI 1.17-1.77, p = 0.0007), lower than the original estimate. This suggests the original dataset may have overestimated the OR, potentially due to selective reporting and small study sizes. Nonetheless, the association remained robust, with diarrhoeic stool showing higher odds of *Campylobacter* detection compared to non-diarrhoeic stool. Heterogeneity remained high after adjustment (I^2^ = 90.52%, p < 0.0001).

Sub-group analysis was run to investigate heterogeneity for OR of *Campylobacter* presence in diarrhoeic stool (Table 2).

**Table 2.** Sub-group analysis of OR for *Campylobacter* presence in diarrhoeic stool versus non-diarrhoeic. * = OR and 95% CI > 1.

| Factor | Number of studies | OR | 95% CI lower | 95% CI upper |
| --- | --- | --- | --- | --- |
| **Study type** |  |  |  |  |
| Case-control * | 34 | 1.98 | 1.57 | 2.49 |
| Cross-sectional * | 17 | 2.04 | 1.42 | 2.93 |
| Cohort * | 9 | 1.66 | 1.01 | 2.72 |
| **Study setting** |  |  |  |  |
| Clinical * | 46 | 2 | 1.66 | 2.42 |
| Community * | 13 | 1.69 | 1.08 | 2.66 |
| Not defined | 1 | 2.03 | 0.08 | 54.83 |
| **Decade** |  |  |  |  |
| 1970 * | 2 | 2.23 | 1.25 | 3.97 |
| 1980 * | 18 | 2.65 | 1.91 | 3.66 |
| 1990 * | 11 | 2.25 | 1.44 | 3.51 |
| 2000 * | 6 | 3.08 | 1.9 | 5.01 |
| 2010 | 18 | 1.27 | 0.97 | 1.64 |
| 2020 * | 5 | 1.3 | 1.01 | 1.68 |

| **Method** |  |  |  |  |
| --- | --- | --- | --- | --- |
| Culture * | 47 | 2.33 | 1.88 | 2.9 |
| PCR * | 11 | 1.19 | 1.03 | 1.39 |
| EIA | 2 | 1.31 | 0.59 | 2.9 |
| Molecular * | 11 | 1.19 | 1.03 | 1.39 |
| Non-molecular * | 49 | 2.25 | 1.82 | 2.78 |
| **Country** |  |  |  |  |
| Egypt * | 5 | 2.3 | 1.7 | 3.12 |
| Democratic Republic of the Congo * | 1 | 2.69 | 1.54 | 4.68 |
| South Africa * | 13 | 2.42 | 1.96 | 3 |
| Rwanda | 3 | 0.94 | 0.68 | 1.29 |
| Ghana * | 1 | 1.41 | 1.06 | 1.86 |
| Tanzania | 6 | 1.06 | 0.82 | 1.37 |
| Malawi * | 2 | 1.66 | 1.27 | 2.18 |
| Guinea-Bissau | 2 | 0.85 | 0.62 | 1.16 |
| Algeria | 1 | 1.22 | 0.79 | 1.87 |
| Zimbabwe * | 2 | 5.45 | 2.24 | 13.25 |
| Nigeria * | 8 | 4.24 | 2.44 | 7.37 |
| Ethiopia * | 3 | 9.57 | 2.61 | 35.12 |
| Madagascar | 3 | 0.96 | 0.7 | 1.33 |
| The Gambia | 2 | 1.46 | 0.26 | 8.26 |
| Burkina Faso | 1 | 2.39 | 0.13 | 43.79 |
| Central African Republic * | 4 | 2.09 | 1.47 | 5.7 |
| The Gambia, Mali, Kenya * | 1 | 1.2 | 1.11 | 1.3 |
| Côte d'Ivoire | 1 | 3.13 | 0.45 | 21.72 |
| Cameroon | 1 | 2.37 | 0.91 | 6.18 |
| **Age Range** |  |  |  |  |
| 0-6 months * | 18 | 2.57 | 1.74 | 3.81 |
| 7-12 * | 7 | 1.6 | 1.07 | 2.4 |
| 13-24 | 10 | 1.02 | 0.68 | 1.52 |
| 25-60 | 7 | 1.76 | 0.77 | 4.05 |
| > 60 | 0 | NA | NA | NA |

Age influenced the odds of detecting *Campylobacter* in diarrhoeic stool compared to non-diarrhoeic stool. Children under 12 months had higher odds of *Campylobacter* detection in diarrhoeic stool (0-6m, OR = 2.57 95% CI: 1.74-3.81; 7-12m, OR = 1.60 95% CI: 1.07-2.40) compared to non-diarrhoeic stool. Children older than 12 months were as likely to have *Campylobacter* detected in diarrhoeic stool as non-diarrhoeic stool (13-24m OR = 1.02 95% CI: 0.68-1.52; 25-60m OR = 1.76 95% CI: 0.77-4.05).

Culture (OR 2.33, 95% CI: 1.88-2.90), and non-molecular methods of detection (OR = 2.25, 95% CI: 1.82-2.78) had higher odds of detecting *Campylobacter* in diarrhoeic stool compared to non-diarrhoeic stool. PCR and molecular methods also suggested higher odds of detection in diarrhoeic compared to non-diarrhoeic stool (OR 1.19 95% CI: 1.03-1.39), but to a lesser degree.

Ten countries showed higher odds of detecting *Campylobacter* in diarrhoeic stool compared to non-diarrhoeic stool, while there was no significant difference in odds for nine countries. Countries in both groups were geographically spread across Africa.

Meta-regression indicated that country had the greatest influence on heterogeneity (R^2^ = 58.14%, I^2^ = 57.62%, p <0.0001), although residual heterogeneity remained. Ethiopia had significantly higher OR (p = 0.0037) for *Campylobacter* presence in diarrhoeic stool. Study setting, detection method and age had some impact on heterogeneity, whilst decade had little effect on heterogeneity (S10).

### Risk factors for *Campylobacter*

Twenty-four studies reported risk factors for *Campylobacter* presence in mixed or diarrhoeic stool using OR. Risk factors evaluated varied widely across studies, with few studies investigating the same risk factor. Further, risk factors were categorised differently, reducing the number of risk factors that could be assessed by meta-analysis. For example, breast feeding was reported as exclusive or mixed, or exclusive until different ages, preventing its inclusion in pooled analysis. Resultantly, pooled crude OR (pCOR) were investigated alongside pooled adjusted OR (pAOR).

Children living in rural, compared to urban, areas had a higher pAOR of detecting *Campylobacter* in diarrhoeic stool (pAOR = 2.59, 95% CI: 1.43-4.69, p = 0.0016). Having contact with animals had a higher pAOR of detecting *Campylobacter* in diarrhoeic stool (pAOR = 4.84, CI: 2.45-9.58, p < 0.0001) than mixed stool (pAOR = 4.28, 95% CI: 2.40-7.61, p < 0.0001), but both were significant. (See Table 3).

**Table 3.**
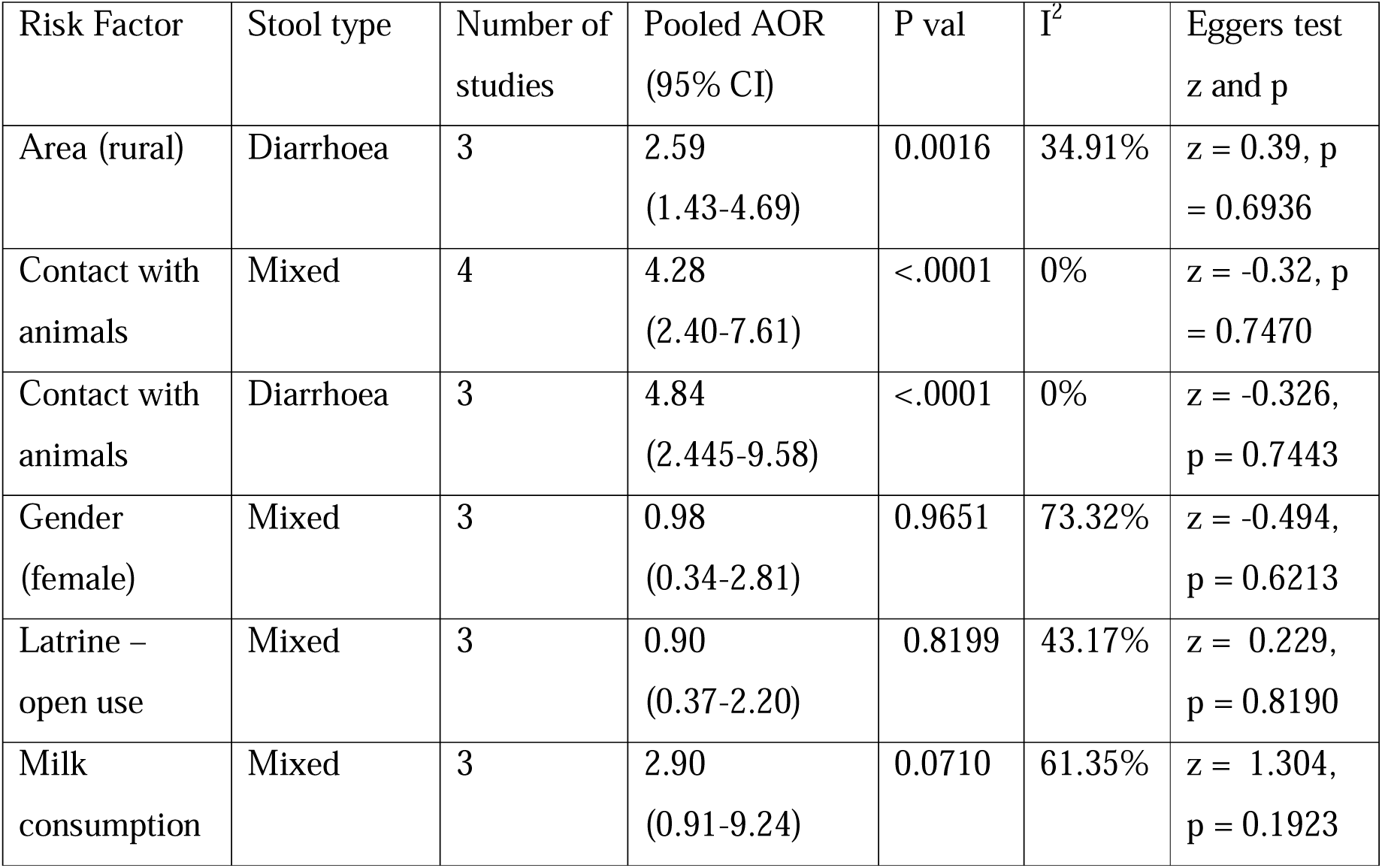
Risk factors for *Campylobacter* presence in stool, pAOR.

Meta-analysis of pCOR identified several potential risk factors for *Campylobacter* presence in stool (S11), this included milk consumption (pCOR = 2.80, 95% CI: 1.21-6.48, p <0.1), not washing hands before feeding children (pCOR = 2.90, 95% CI 1.33-6.29, p < 0.01), access to unsafe water (pCOR = 1.97, 95% CI: 1.22-3.17, p < 0.01) and vomiting (pCOR = 2.77, 95% CI: 1.86-4.13, p < 0.0001). Unsafe water consumption had a pCOR of 3.31 (95% CI: 1.04-10.48, p < 0.05) for diarrhoeic stools. Latrine use in diarrhoeic stools had a significant OR after removing one influential study (pCOR = 2.73, 95% CI: 1.55-4.81, p < 0.0005).

### AMR

AMR data for antibiotics of interest were reported in 40 studies, of which four studies used multiple methods of determining AMR. Disk diffusion was used in 34 studies (85.00%), E-tests were used in five studies (12.50%), two studies used agar dilution (5.00%), two studies detected AMR genes (5.00%) and one study (2.50%) did not report the method used. Thirteen studies used CLSI and four studies used EUCAST international standards for determining AMR. Studies came from 12 countries, with Ethiopia (n=9), Nigeria (n=8) and Egypt (n=7) contributing the most studies (S12).

The prevalence of *Campylobacter* AMR was lowest for chloramphenicol (9.85%, 95% CI: 0.79-24.96%) and highest against streptomycin (48.94%, 95% CI: 20.17-78.07%). Significant heterogeneity existed for each antibiotic (I^2^ > 80%), other than doxycycline (I^2^ = 39.84%) (Table 4).

**Table 4.**
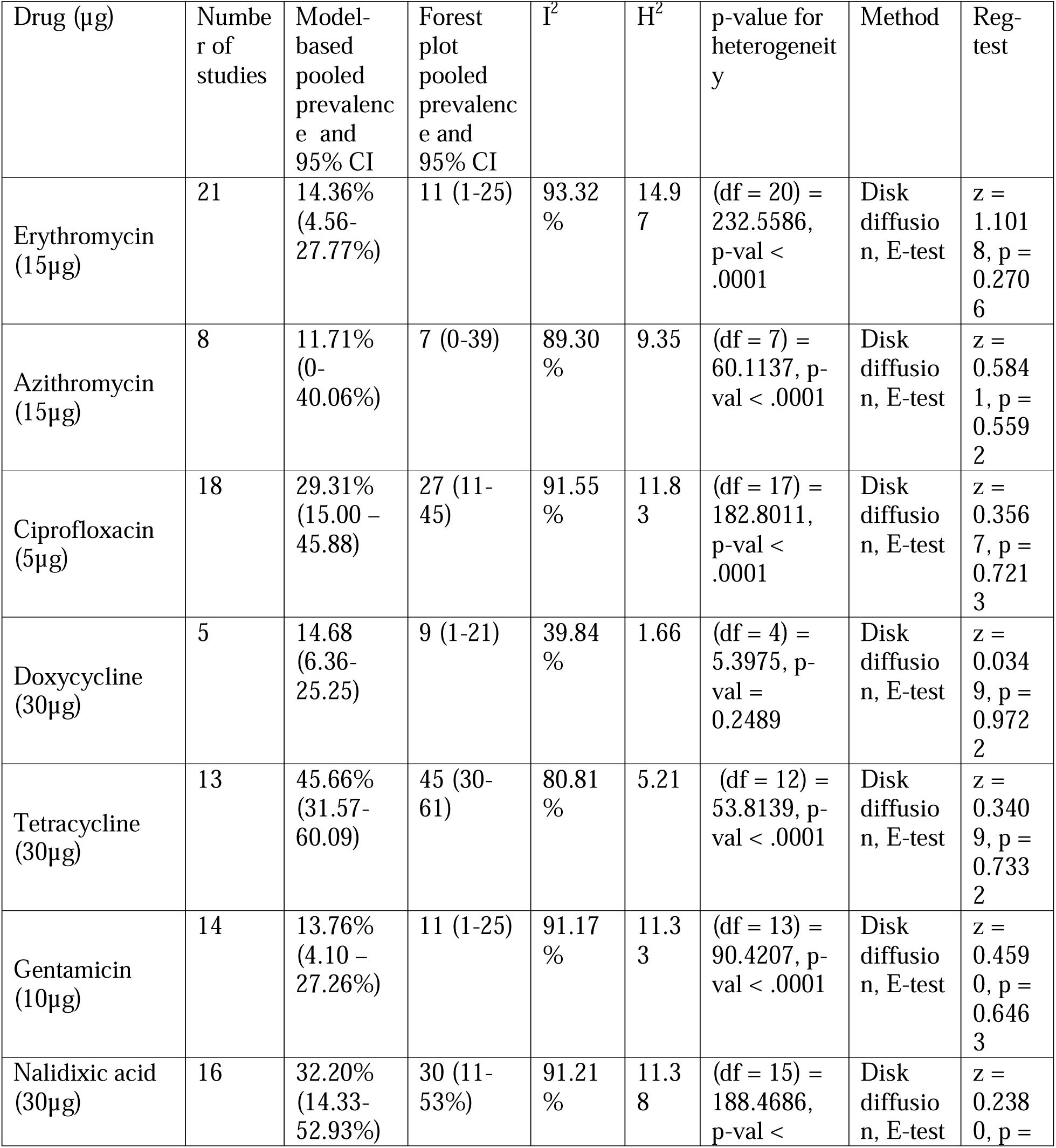
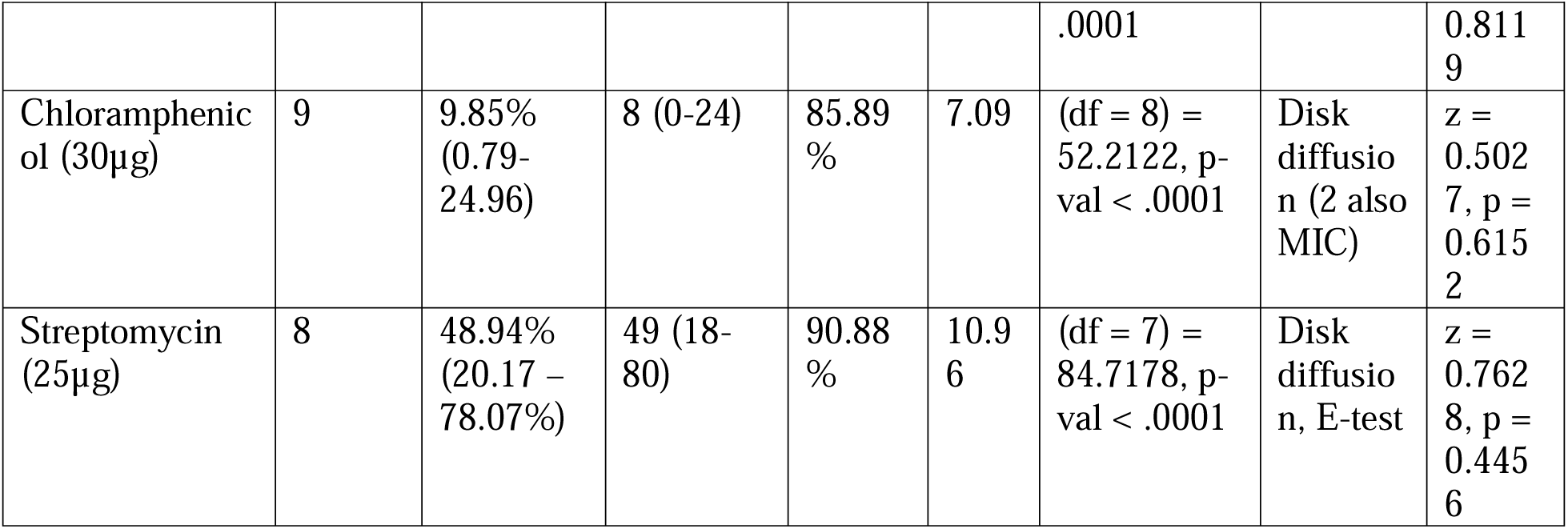
Pooled prevalence of AMR in *Campylobacter* µg in brackets represents the concentration of drug assessed where disk diffusion method was used.

Meta-regression identified that AMR prevalence was significantly higher in the decade 2000 for streptomycin and 2020 for erythromycin, doxycycline, tetracycline, nalidixic acid and streptomycin. Resistance was not affected by decade for the remaining antibiotics. This may suggest a generalised increase in AMR from 2020 onwards, though this should be interpreted with caution. The amount of heterogeneity accounted for by country varied widely from 100% for azithromycin (R^2^ = 100%) to 0% for doxycycline and streptomycin (R^2^ = 0%). Studies in Egypt had a positive influence on AMR resistance (S13).

## DISCUSSION

This meta-analysis estimated the pooled prevalence of *Campylobacter* in children, differentiating between stool type to quantify the relative significance of *Campylobacter* in diarrhoeic and asymptomatic children. Included studies investigated *Campylobacter* presence across a wide age range, from birth up to 18 years old. However, the importance of diarrhoea in children under 5 years (4) is reflected, with over two thirds of studies focusing on *Campylobacter* in this age group.

This review suggests *Campylobacter* prevalence in diarrhoeic children may be higher across Africa (11.45%, 95% CI 5.38-8.72%) than South Asia, where the pooled prevalence was 6.96% (95% CI 5.38-8.72%) (226). Our findings are consistent with a Sub-Saharan Africa review of *Campylobacter* prevalence in people across all age groups (18). They detected a pooled prevalence of *Campylobacter* of 9.9% (95% CI 8.4-11.6%) with no significant difference between diarrhoeic and non-diarrhoeic individuals (18). Our review detected a similar prevalence of *Campylobacter* in diarrhoeic and non-diarrhoeic stool (11.45%, 95% CI 5.38-8.72% and 12.82%, 95% CI 7.81-18.82% respectively), while the prevalence was double this (31.26%, 95% CI 17.34-47.17%) in mixed stool types. It is postulated that the method of detection influenced the high pooled prevalence in mixed stool types, where method contributed significantly towards heterogeneity. An equal number of studies detected *Campylobacter* via PCR and culture, with PCR methods giving a pooled prevalence over double that of culture. A significant difference in pooled prevalence was seen between culture and PCR for non-diarrhoeic studies (7.81%, 95% CI 4.04-11.11% and 30.14%, 95% CI 16.01-46.53% respectively) but was minimal for diarrhoeic stool (culture = 10.91%, 95% CI 8.98-13.01%, PCR = 10.89%, 95% CI, 6.21-16.67%). In both categories the number of studies using PCR was, at most, a quarter of all studies contributing to the analysis. Therefore, the relative contribution of PCR studies may have increased the pooled prevalence of *Campylobacter* in mixed stool. Further, different culture and PCR techniques vary in their sensitivity and specificity for *Campylobacter* detection. Consequently, it will be important to continue monitoring the prevalence of *Campylobacter* in diarrhoeic and non-diarrhoeic stool if PCR methods become more common place.

Prevalence was similar across all stool types for *C. jejuni* and *C. coli*, with diarrhoeic stools being slightly higher than non-diarrhoeic and mixed stool types. *C. jejuni* had more than double the prevalence of *C. coli,* with the pooled prevalence of *C. jejuni* in diarrhoeic stool (14.63%, 95% CI: 9.68-20.39%) being greater than the pooled prevalence of all *Campylobacter* spp. (11.45%, 95% CI: 5.38-8.72%). This is not surprising, as *C. jejuni* is the species most frequently associated with diarrhoea. Additionally, culture methods may be biased towards detecting *C. jejuni* and *C. coli*. The introduction of metagenomic sequencing has revealed a higher diversity of *Campylobacter* species in human stools, including multiple species from the same participant (138). This diversity may help explain some of the differences in prevalence. *C. jejuni* and *C. coli* may account for a greater proportion of *Campylobacter* species detected in diarrhoeic stools, and other *Campylobacter* species account for a greater proportion in non-diarrhoeic or mixed stool.

This review highlights the high prevalence of asymptomatic *Campylobacter* species carriage. While this has been suggested to adversely affect child development (14, 27), its epidemiological burden and developmental consequences in African children remain poorly characterized. This substantial prevalence underscores the need for further investigation into the clinical and developmental impact of *Campylobacter* carriage in this high-burden population.

The odds of detecting *Campylobacter* in diarrhoeic stool were greater than in non-diarrhoeic stool, despite similar pooled prevalence between diarrhoeic and non-diarrhoeic stool. Prevalence reflects averages across studies, while ORs describe within study associations, accounting for study-level differences. Thus, ORs offer a more appropriate measure for determining the association between *Campylobacter* and diarrhoea. The strength of association was age dependent. *Campylobacter* was more likely to be detected in diarrhoeic stool of children under 12 months, with the strongest association in those under six months. Above 12 months, no association between *Campylobacter* presence and stool type was observed. In endemic settings, children are exposed to *Campylobacter* from a young age, with initial exposure often leading to diarrhoea. As immunity develops through repeated exposed, children may acquire partially protective immunity (11) and carry *Campylobacter* asymptomatically. The age at which the switch between symptomatic and asymptomatic *Campylobacter* carriage occurs is not clearly defined. *Campylobacter* prevalence in stool increases up to 12 months (227), and over 90% of children are seropositive for antibodies against *Campylobacter* (110, 210) by this age. Together with the findings of this meta-analysis, this suggests that in the African setting diarrhoea is mostly associated with children up to one year of age. More research into the development of immunity and the effect of subclinical carriage is warranted.

Contact with animals was identified as a risk factor for *Campylobacter* presence in mixed and diarrhoeic stools. Chickens have often been implicated as risk factors for *Campylobacter* presence in human stool (228, 229). However, we did not identify any species of animal as a risk factor at pCOR nor pAOR, the latter due to insufficient data. Therefore, limiting children’s contact with any animals or their environment may reduce the odds of exposure to *Campylobacter*.

Children in rural areas were more likely to have *Campylobacter* in their stools than in urban areas. All studies that assessed this risk factor were on diarrhoeic stool, therefore it is not possible to state if being in a rural area is a risk factor for carriage of *Campylobacter* regardless of stool type.

AMR was greatest against streptomycin and tetracycline, which are infrequently used to treat *Campylobacter* in children. However, they have been widely used in livestock (230) suggesting spillover of bacteria and resistance from animals to children in Africa. Resistance to fluoroquinolones (ciprofloxacin), one of the first line antibiotics for *Campylobacter*, was high. In comparison, first line antibiotics belonging to the macrolide class, azithromycin and erythromycin, had low resistance prevalence, suggesting their use remains effective in Africa. However, low availability of these drugs, as has been demonstrated in Malawi and Tanzania (231–233), may limit their use.

*C. jejuni* isolates from Peruvian children under five years old had a broadly similar resistance profile (234) to *Campylobacter* in these studies. Slightly lower levels of resistance were identified in isolates from Peru for erythromycin (5.3%), azithromycin (4.9%), chloramphenicol (0.2%) and gentamycin (1%), whilst tetracycline (55.8%) resistance was greater. Across Africa resistance to ciprofloxacin and nalidixic acid was about half that reported in Peru (77.4% and 64.9%, respectively). These geographic variations underscore the critical need for coordinated region-specific AMR surveillance to guide evidence-based treatment recommendations.

Assessment of risk factors of *Campylobacter* presence in stool had limitations. Comparatively few studies investigated risk factors for *Campylobacter*. Further, there was a lack of consistency across studies in the type of risk factors investigated, and how they were characterised. This meant that some risk-factors could not be analysed despite being investigated in multiple papers. To address this, we recommend that a standardized set of risk factors should be defined. Moreover, risk factors for *Campylobacter* presence were identified in mixed or diarrhoeic stool types, and pooled OR calculations may have involved different stool types. Risk factors may differ for *Campylobacter* presence in mixed stool types versus diarrhoeic stool. For example, in this review the pCOR’s suggested that latrine use affected the likelihood of *Campylobacter* presence in both mixed and diarrhoeic stool. No association was found when calculating the pAOR of mixed stool types. Not enough studies reported aOR for the significance of latrine use on diarrhoeic stool to be assessed. Further investigation into risk factors for *Campylobacter* presence in diarrhoeic stool is warranted. Additionally, no studies identified risk factors in non-diarrhoeic stool, preventing associations between these risk factors and those for diarrhoeic stool.

Heterogeneity was high across all analyses, likely due to the broad scope of this review. This review assessed prevalence, association with diarrhoea, risk factors and AMR across Africa. Resultantly, a wide selection of papers were included. Heterogeneity is to be expected for a review across a continent, and country frequently contributed towards heterogeneity. Despite this the meta-analysis may have missed some non-English language studies, and therefore additional country data, which may have biased findings. The review spanned 6 decades, potentially contributing to heterogeneity. The prevalence of *Campylobacter* in diarrhoeic stool varied little between decade (S9). However, there was an increasing trend in *Campylobacter* prevalence in both non-diarrhoeic and mixed stool types between 1990 and 2020. The introduction of advanced detection methods, such as PCR, in recent decades may have influenced prevalence calculations. Further, multiple methods of culturing *Campylobacter* (see S14 for details) may contribute towards heterogeneity.

## CONCLUSIONS

This review found high and comparable *Campylobacter* prevalence between diarrhoeic and non-diarrhoeic stool, highlighting the substantial burden of asymptomatic carriage. Further research into the risk factors for and impacts of asymptomatic carriage is warranted. *Campylobacter* presence in stool of children under the age of 12 months is more likely to be associated with diarrhoea. In children above this age there was no association between *Campylobacter* presence and stool type. This may be due to age related immunity and an associated shift from infection to asymptomatic carriage.

Contact with animals and living in a rural environment were consistently identified as risk factors for *Campylobacter* presence in stool. Standardising risk factor assessment across future studies would enable effective comparisons and provide evidence of transmission pathways specific to African populations, on which future control messages can be targeted.

AMR analysis found that *Campylobacter* remains largely susceptible to first line macrolide antibiotics. Resistance is developing against antibiotics typically associated with use in livestock, suggesting zoonotic transmission. Continued AMR monitoring is essential to track resistance trends, preserve treatment efficacy and inform empirical treatment guidelines.

## Supporting information

Prisma Checklist

Prisma abstract checklist

## Data Availability

All data produced in the present study are available upon reasonable request to the authors

## Acknowledgements

We would like to thank Priscilla Dzanja for her contribution in the initial screening of titles and abstracts for this systematic review.

