## Supplementary material for "The Prevalence, Risk Factors, and Antimicrobial Resistance of *Campylobacter* in African Children: A Systematic Review and Meta-Analysis": S14

### Culturing *Campylobacter*

##### Culturing methods

Studies reported a range of methods for culturing *Campylobacter*, including a variety of media and methods of generating microaerophilic environments. Skirrow media (n = 25) was used most frequently, followed by Butzler (n = 19), then mCCDA (n = 19). Some studies used multiple media, and some compared the ability of the different media to isolate *Campylobacter*. The most common media combination was Skirrow and Butzler (n = 5). Studies that used a *Campylobacter* selective media were grouped together. Studies that stated a selective media without specifying it as selective for *Campylobacter* were grouped together separately (Table 1).

| **Media** | **Count** |
| --- | --- |
| Skirrow | 25 |
| Butzler | 19 |
| *Of which Skirrow and Butzler* | *5* |
| mCCDA | 19 |
| Modified Skirrow | 5 |
| Preston | 6 |
| Karmali | 6 |
| Cape Town | 4 |
| Butzler type | 3 |
| CHROMagar | 4 |
| *Campylobacter* selective (incl CCDA) | 17 |
| Selective media | 6 |
| Standard | 6 |
| Not described | 5 |
| Other | 22 |

Table 1. Media used to culture *Campylobacter*

An enrichment broth was used by 20 studies, of which six used Bolton broth, six Preston broth, two thioglycolate broth, and one each for Butzler, Muller-Hinton and TSB. Two papers described the broth as *Campylobacter* enrichment broth. One paper did not describe the type of broth.

*Campylobacter* requires a microaerophilic environment for growth. Gas generating methods were used by 44 studies of which 16 used Campygen, 3 used BBL and 2 used Anaeropacks. Candles were used in 19 studies. Many studies (n = 31) stated that a microaerophilic environment was generated but did not state how, whilst others did not reference a microaerophilic atmosphere (Table 2).

| **Atmospheric condition** | **Count** |
| --- | --- |
| Gas generating | 44 |
| *Of which Campygen* | *16* |
| *Of which BBL* | *3* |
| *Of which Anaeropack* | *2* |
| Microaerophilic | 31 |
| Candle | 19 |
| Not described | 16 |
| Standard methods | 12 |
| Referred to culture method or another paper | 4 |
| Anaerobic | 1 |
| Jar | 1 |
| Gas Mixture | 1 |

Table 2. Methods described for creating a microaerophilic environment.

##### Discussion

The fastidious nature of *Campylobacter*, requiring specific conditions and specialised equipment for growth, make culture challenging. Generating a microaerophilic environment is essential for successful culture. This can be reliably created using variable atmospheric incubators, which are expensive. None of the studies included in this review reported using this method, with most using gas packs that absorb oxygen and increase carbon dioxide. No consensus exists for the preferred culture methodology, with internationally recognised quality assurance schemes recommending an array of protocols (1). Protocols differ in their ability to grow *Campylobacter* (2). Seven methods of generating a microaerophilic environment and over 13 types of media were described in this review, likely contributing towards heterogeneity. Whilst some heterogeneity could be accounted for, residual heterogeneity frequently remained. This may reflect true differences between studies and, or publication and small study bias.

##### *A note on Campylobacter detection methods in Africa*

As noted in the main paper, culture or non-molecular methods were more likely to detect *Campylobacter* in diarrhoeic stool than non-diarrhoeic stool, whereas molecular methods showed consistent detection across stool types. The higher sensitivity of molecular methods allows detection of lower bacterial loads that culture-based approaches may miss (3). This review found that molecular methods identified a higher overall prevalence of *Campylobacter* than non-molecular methods across mixed, non-diarrhoeic and diarrhoeic stool types.

This finding has important implications for resource limited settings, particularly Africa, where molecular methods are not widely available. Over 70% of studies in this review used culture-based detection. The widespread use of culture in the region reflects practical constraints; molecular methods remain inaccessible in many African countries due to high cost and infrastructure limitations. Previous studies report an association between *Campylobacter* load and diarrhoea severity, suggesting culture may under detect *Campylobacter* with low bacterial load (4). As such, *Campylobacter* carriage and diarrhoea may be underestimated in regions without access to PCR-base diagnostics, potentially masking the true burden of *Campylobacter* in Africa.
