## Supplementary figures and images for "The Prevalence, Risk Factors, and Antimicrobial Resistance of *Campylobacter* in African Children: A Systematic Review and Meta-Analysis"

### Figure S9

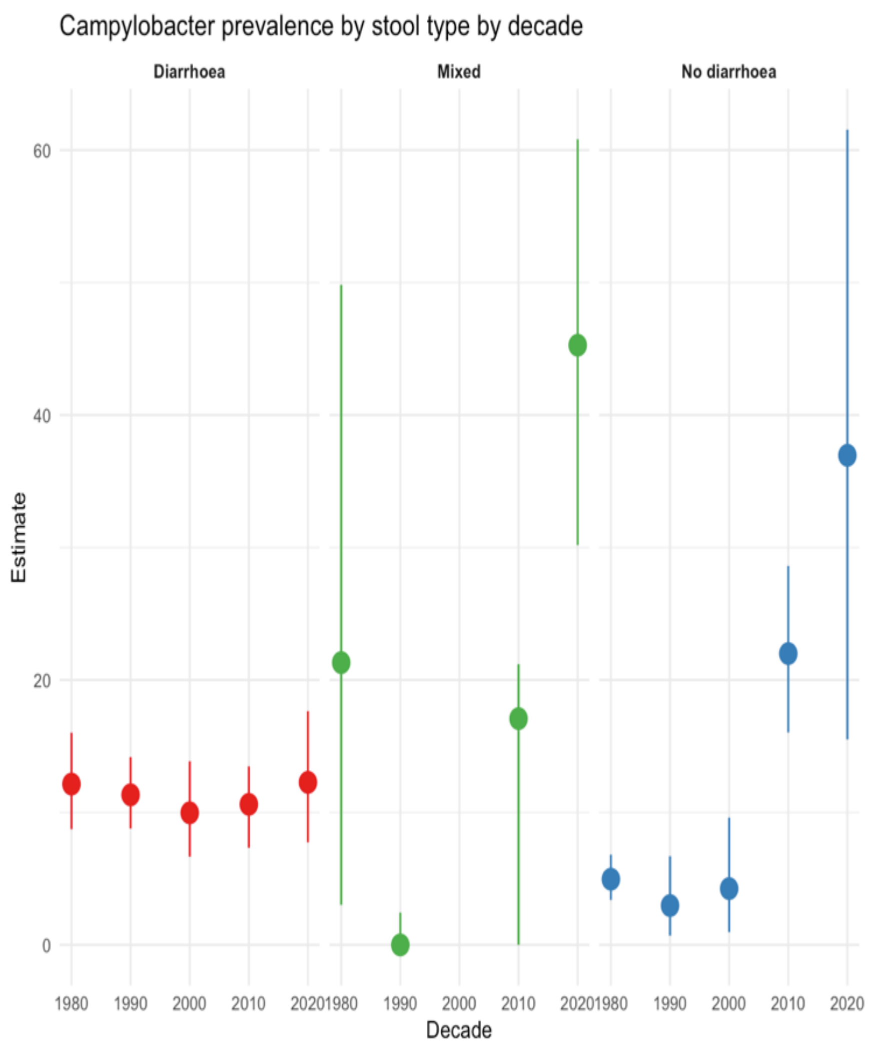


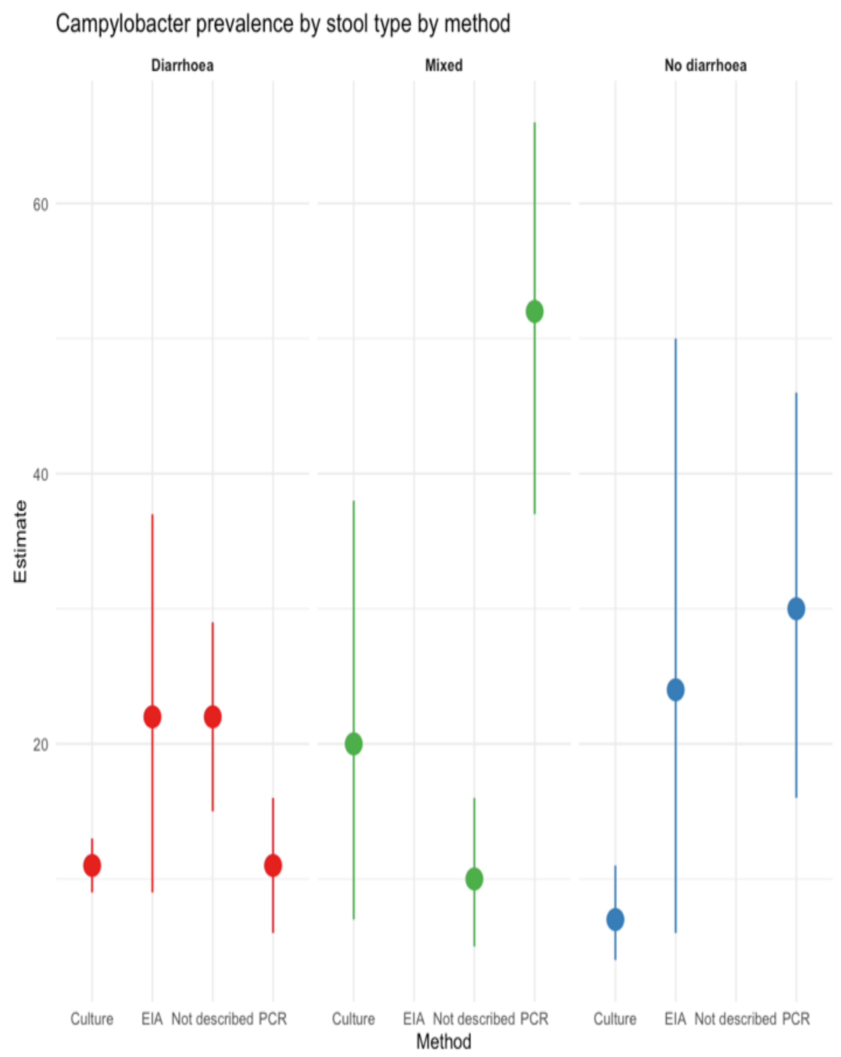


Figure S9. Prevalence in stool type by decade and method of detection.
